# Human Leukocyte Antigen-C and Haplotypes: Associations with Resistance and Susceptibility to HIV-1 Infection among Serodiscordant Couples in Nigeria

**DOI:** 10.1101/2021.04.15.21255498

**Authors:** Ngozi Mirabel Otuonye, Ma Luo, Onaiwu Idahosa Enabulele, Ayorinde James, Felix Emele, Prince Ele, Thomas Bielawny, Mercy Mayowa Ojetunde, Nkiruka Nnonyelum Odunukwe

## Abstract

**INTRODUCTION:** The Human Leucocyte Antigen (HLA) class-1 is known to play a significant role in mediating resistance or susceptibility to HIV infection in the clinical course of AIDS. Recent studies have identified HLA-C as a key molecule that affects HIV disease progression. However, the role of HLA class 1 in heterosexual HIV-1 susceptibility or resistance in serodiscordant couples is not known in Nigeria. Therefore, this study evaluated the association between HLA-C susceptibility and resistance in HIV-1 transmission amongst heterosexual serodiscordant couples in Nigeria.

**METHODS:** A total of 271 serodiscordant, concordant HIV positive and negative couples who gave informed consent were enrolled into this study. Extracted genomic DNA was sequenced for high resolution HLA-C class 1 genotypes using allele-specific primers (on exons 2 and 3) for HLA-C sequencing and typing.

**RESULTS:** The highest frequency distribution of high-resolution HLA-C alleles observed in the HIV positive subjects were: HLA-C*040101 178 (35.0%) followed by C*0701 124 (24.9%) compared with HIV negative subjects: C*040101 108(39.0%) followed by C*0701 64(24.7%). Alleles C*070201 (OR = 4.19, *P<0*.*05*) and C*0804 (OR = 3, *P*<0.045) were found to be independently associated with HIV-1 susceptibility in the cohort of serodiscordant couples. HLA-C*0802 (OR=0.5. *P*<0.005) and C*0304 (OR=0.34. *P*<0.002) were significantly associated with HIV-1 resistance to HIV-1 infection among the cohort.

**CONCLUSION:** The result has contributed to the importance of how host HLA-C genetic factors can influence HIV-1 disease susceptibility (HLA-C*070201; C*0804) and resistance (HLA-C*0802; C*0304) in serodiscordant couples. This information may contribute to the development of future effective HIV vaccine in Nigeria.

## INTRODUCTION

Human Immunodeficiency Virus (HIV) infection remains one of the greatest scourges of our time (Kaiser, 2017 and UNAIDS, 2019), particularly in Sub-Saharan Africa – with over 65% of the world’s HIV infection. Nigeria is considered to have the second largest HIV epidemics in sub-Saharan Africa, and one of the highest rates of new HIV infection in sub-Saharan Africa (UNAID, 2019). Heterosexual exposure is the primary mode of transmission in Nigeria and involves mainly HIV-1. An estimated 70% of HIV-1 infection occurs between married partners and co-habitation with HIV infected person represents the largest HIV risk group (Bagala, 2006). Evidences indicate that significant number of HIV-1 infected couples in Nigeria are in serodiscordant relationship (Nnebue *et al*., 2017). Serodiscordant relationship is a term used to describe a couple in which one partner is infected by HIV and the other is not. Studies have implicated the HLA as important host immune response to HIV-1 infection and also suggest that the make-up of a person’s HLA affects the individual’s HIV disease progression (Tang *et al*., 2008). The HLA are genetically inherited protein receptors that are present on the surface of human cells and involved mainly in self and non - self recognition of antigenic peptides. The HLA is the human version of the Major Histocompatibility Complex and is located at the short arm of human chromosome 6 (6p21), which spans approximately 4.0 kilobases of DNA, and contains more than 220 genes of diverse functions (Choo, 2007). The HLA is known to be the most polymorphic genetic system in humans and made up of class-I (HLA-A, -B and -C) and class-II (DP, DQ, DR, DM, DP and DP) (Tang *et al*., 2008; Lingappa *et al*., 2008).

The biological role of HLA class I and II molecules are to present processed antigenic peptides to CD8+ and CD4+ T lymphocytes respectively for immune recognition and stimulation, with the aim of destroying the pathogen from which the antigenic peptide came (Choo, 2007). The MHC-1 molecules present viral peptides to CD8+ T-cells and serve as ligands for killer cell immunoglobulin-like receptors (KIRs). A sub-set of MHC-1 allotypes are associated with delayed disease progression and effective control of viral replication, most notably HLA-B 5701 (Kulpar *et al*., 2011).

Previous study by Fellay *et al*. (2011) identified HLA-C as a key molecule that affects HIV disease progression. In the event of HIV-1 infection, HLA-A, -B and -C proteins can all present viral peptides which are recognized by CD8+ T cells (Bachtel *et al*., 2018). However, HLA-C allotypes dominate over HLA-A and HLA-B allotypes as a source of ligands for KIRs (Aguilar *et al*., 2010). The HLA-C plays a crucial role as a molecule capable of sending inhibitory signals to both natural killer (NK) cells and cytotoxic T lymphocytes (CTL) via binding to the killer cell IgG-like receptors. The tendency of HLA-C to be expressed as open conformer, upon cell activation, endows it with unique capacity to associate with other cell surface molecules as well as with HIV-1 protein, Zipeto and Beretta, (2012). Furthermore, promoter alleles express higher levels of HLA-C on T-lymphocyte that have significant control of HIV-1 viraemia and progress more slowly to AIDS. The Negative Regulatory Factor (NEF) protein also disrupts antigen presentation in association with HLA -A and HLA-B, but not HLA-C. This shows that HLA-C efficiently presents antigens to CTLs in HIV infection for immune recognition and stimulation.

A study on HIV-1 discordant couples in Zambia showed that viral transmission from index case to seronegative partners can vary according to specific HLA class I alleles or haplotypes (Tang *et al*., 2018). The role HLA plays in HIV-1 transmission in serodiscordant couples in Nigeria is unknown. Data on HLA-C alleles associated with HIV-1 transmission in serodiscordant relationship is lacking in Nigeria. Therefore, this study aimed at providing some information in this respect.

Therefore, the objectives of this study are to:

1. Document the frequencies of HLA-C alleles in HIV positive and negative individuals
2. Determine the role HLA-C class 1 variants play in HIV-1 resistance and or susceptibility to HIV-1 infection among serodiscordant couples.
3. Document the profile of Long-Term Non-Progressors’ HLA-C alleles, baseline and follow-up of CD4 count and Plasma viral load (HIV-RNA) among the index and partners.

## MATERIALS AND METHODS

### Study Population

The study population comprised HIV-1 serodiscordant couples attending the HIV clinics of Nnamdi Azikiwe University Teaching Hospital (NAUTH), Nnewi Nigeria, and Nigerian Institute of Medical Research (NIMR), Yaba, Lagos. HIV negative couples used as controls were recruited among NIMR Workers (NIMRW) and Divine Grace Church (DGC).

### Inclusion Criteria

Couples who have discordant HIV test results, co-habited for at least 3 months, have more than one partner, are 16-50 years of age (for female) or 16-65 years (for male), and HIV positive partner is or not on ARV.

### Exclusion Criteria

Couples who are not within the required age group and have not co-habited for up to three months.

### Study Sites

Samples were collected and processed in two government-owned health institutions namely: Nigerian Institute of Medical Research (NIMR), Yaba, Lagos Nigeria, and Nnamdi Azikiwe University Teaching Hospital (NAUTH), Nnewi, Anambra State, Nigeria. Molecular experiments were carried out at National Microbiology Laboratory, Winnipeg, Manitoba, Canada.

### Study design

Case–control study of 271 couples: people living with HIV/AIDS and their HIV negative partners (serodiscordant couples), concordant HIV positive couples and concordant HIV negative couples (control) were recruited. The index may or may not be on Antiretroviral.

### Focused group discussion

Participants were counselled and told to inform their partners about the research - to facilitate their consent. The partners of all index were screened for HIV. Those who subsequently turned out to be HIV positive were referred to NIMR and NAUTH HIV clinic for management. Couples recruited from NIMR workers (NIMRWC) and Divine Grace Church Couples (DGCC) were screened for HIV to confirm their sero-negative status.

Structured questionnaires were administered to the couples to obtain information on socio-demographic characteristics, knowledge of partners’ HIV status, knowledge of STIs, laboratory tests and treatment status, sexual behaviours and practices, were required for the completion of the questionnaire.

### Methods of Sample Collection and DNA Extraction

A total of 542 blood samples were collected from 236 couples from NIMR, and 35 couples from NAUTH. Whole blood was collected (by vein puncture) in EDTA-containing bottles for HIV antibody detection. The white cell pellet was separated from buffy coat by centrifugation of the blood samples at 2000 rpm for 15 minutes. DNA extraction was done using gnomic DNAQIAGEN kit as described by the manufacturers. Extracted DNA was placed in the cryovial and stored frozen at −80^0^C for further analysis.

### Laboratory Investigations

#### DNA Amplification

Amplification of the DNA was made by using Polymerase Chain Reaction (PCR). Each PCR reaction was composed of 20µl of ddH2O, 5µl of gDNA, and 25µl of master mix. The master mix contained 22.75µl of 2X mix (120mM Tris-HCl, 3mM MgCl2, 30mM (NH4)2SO4, 200µM dNTPs, 0.2% gelatin, ddH2O), 55 pmol of both forward and reverse primers (Primer structural sequence is as shown below) and 13.75 µl of Taq polymerase. For HLA -C typing, primers were set to detect polymorphisms in exons 2 and 3 of the class 1 genes as these regions cover most of the know polymorphisms. The structural sequence of the primers used for HLA-C was CPCRR (representing HLA-C Reverse primers), and CPCRF (representing HLA-C Forward primers), as shown below.

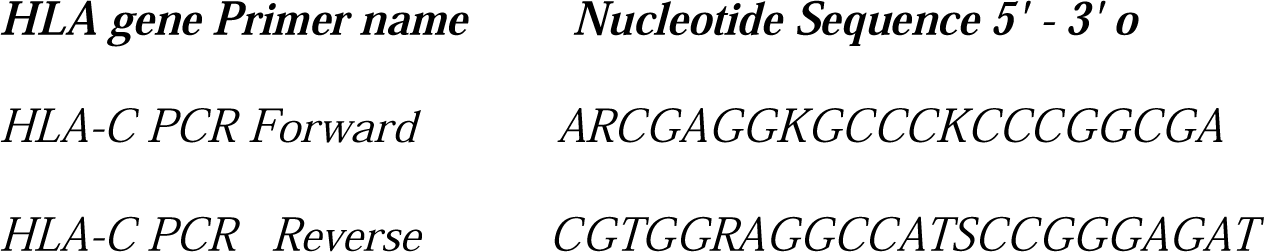

The PCR program was as follows: a 5 minute initial denaturation at 96^°^C, followed by 44 cycles of one minute of denaturation at 96^°^C, one minute of primer annealing at 5^°^C below the Tm of the respective primers and two minutes at 72^0^C for elongation. Each PCR programme finished with a final elongation step at 72^°^C for ten minutes. After amplification, Agarose Gel Electrophoresis (1% Agarose gel) was used to identify the amplified DNA products

### Purification of PCR Product

Following the PCR, the product was precipitated using 21µl of a mixture containing 5ml of 95% ethanol and 250µl of sodium acetate. Finally, the content of each well was transferred to MicroAMP™ plates and put in the Applied Biosystems™ 3130xl Genetic Analyzer for electrophoresis, according to Agencourt® AMPure Protocol (2007).

### Sequencing HLA PCR, using Sanger Sequencing Method (Sanger *et al*., 1997)

This was performed after the PCR products were purified. Sequencing was focused on the most polymorphic exons of the class 1 genes. To genotype HLA-C alleles, exons 2 and 3 were sequenced (21). Each sequencing PCR reaction contained 4µl of purified PCR product, 1.5µl primer at 3.2µM and 2µl of Applied Biosystems™. BigDye® Terminator V1.1 and the forward and reverse primers as shown on the table 1 below:

**Table 1.** Sociodemographic Characteristics of Study Participant (n = 542)

| Sites: NIMR/NAUTH |  | Concordant HIV Positive |  | Concordant HIV Negative |  | Serodiscordant Couples |  |
| --- | --- | --- | --- | --- | --- | --- | --- |
| Age group | Total no - 52 | Frequency (%) | Total no -42 | Frequency (%) | Total no - 448 | Frequency (%) | Total |
| **Age** |  |  |  |  |  |  |  |
| 20-25 | 2 | (0.3) | 0 | (0.0) | 7 | (1.3) |  |
| 26-30 | 2 | (0.4) | 2 | (0.4) | 56 | (10.3) |  |
| 31-35 | 10 | (1.9) | 6 | (1.1) | 114 | (21.0) |  |
| 36-40 | 14 | (2.6) | 10 | (1.9) | 118 | (21.9) |  |
| 41-45 | 12 | (2.2) | 12 | (2.2) | 76 | (14.0) |  |
| 46-50 | 4 | (0.73) | 7 | (1.3) | 46 | (8.5) |  |
| >50 | 8 | (1.5) | 5 | (0.9) | 31 | (5.8) |  |
| **Total** | **52(26)** | **(9.5)** | **42(21)** | **(7.8)** | **448 (224)** | **(82.7)** |  |
| **Sex** |  |  |  |  |  |  |  |
| Male | 26 | (4.7) | 18 | (3.32) | 224 | (41.35) | **50.47** |
| Female | 26 | (4.7) | 24 | (4.42) | 224 | (41.35) | **49.37** |
| **Marital Status** |  |  |  |  |  |  |  |
| Engaged | 0 | (0.0) | 0 | (0.0) | 16 | (3.0) | **3.0** |
| Married | 52 | (9.54) | 42 | (7.8) | 432 | (79.7) | **97.0** |
| **Ethnic Groups** |  |  |  |  |  |  |  |
| Ibo | 36 | (6.6) | 23 | (4.2) | 239 | (44.1) | **54.9** |
| Yoruba | 15 | (2.8) | 10 | (1.8) | 87 | (16.1) | **20.9** |
| Hausa | 0 | (0.0) | 3 | (0.6) | 47 | (8.7) | **9.3** |
| Others | 8 | (1.5) | 2 | (0.4) | 73 | (13.0) | **14.9** |
| **Total** | **542** | **(100)** |  |  |  |  |  |

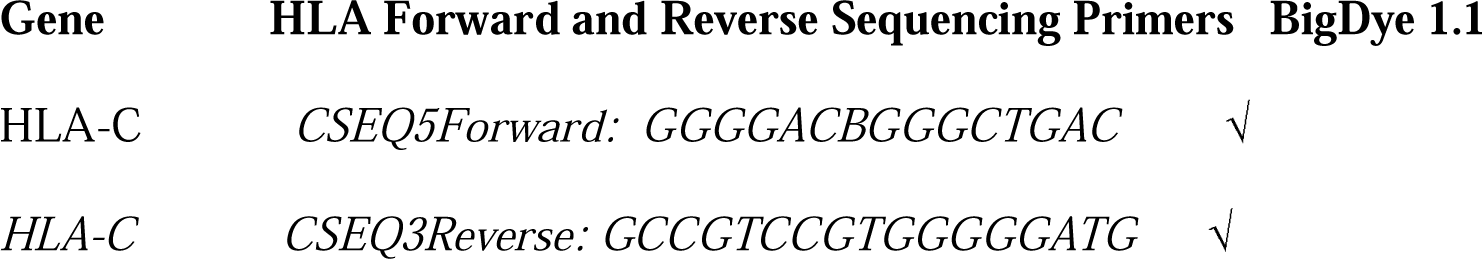

For sequencing-PCR, each primer (forward or reverse) was in separate reactions.

### Method

The BigDye v1.1 and primers were defrosted in a tray of ice.

Using a Pipette, **44.5** µl of the BigDye and Primer was added to the **first column** of the plate, **3.5** µl of master mix was pipetted to the remaining columns, and **4**μ**l DNA** was added to each column. Plates were sealed with silicone foil, quick spun and placed in thermocycler for 8 hrs. Each PCR master plate of 96-wells was done twice. One 96 -well plate contained reverse primers and another 96 -well plate contained forward primers. During the elongation steps, deoxynucleoside triphosphates (dNTPs) and smaller amounts of fluorescently labelled dideoxynucleoside triphosphates (ddNTPs) were added into the growing strand. The sequencing PCR program commenced with an initial denaturation at 96°C for three minutes, followed by 80 cycles of denaturation at 96°C for thirty seconds, primer annealing at 50°C below its Tm for thirty seconds, and finally an elongation step at 60°C for four minutes. A total of 8 hrs was spent to assay each HLA PCR Sequencing reaction.

### Ethanol Precipitation

Following Sequencing PCR, the PCR plate was quick spun and 21µl of a mixture containing 5ml of 95% ethanol and 250µl of sodium acetate was dispensed into the plates. This process removed any buffers, salts and reagents in the PCR product. The plate was sealed with a metal foil, vortexed and spun quickly. The plate was then placed in a −20°C for 1 hour. Plate was spun at 4000rpm for I hour (Long spin step). Finally, the contents of each well were transferred to MicroAMP™ plates and put in the Applied Biosystems™ 3130xl Genetic Analyzer for electrophoresis.

### Biosystems™ 3130xl Genetic Electropherogram

Each participant’s sequenced PCR product was loaded in the Biosystems for electrophoresis and data analysed, using appropriate software, and interpreted according to Applied Biosystem manual.

### Condon Express TM and Sequencher V5 for Data Integrity

CodonExpress was used to type the Sequence-Based HLA class 1 genes as described by the University of Manitoba, Canada instruction manual (2010). Data Integrity was done by using DNA Sequencer V5 Software to rule out all unresolved HLA typing ambiguities.

### Statistical Analysis

Data generated were entered into IBM SPSS statistics version 20. Cross-sectional analysis using Chi Square was used to identify association between genetic and non-genetic factors. Confidence interval was set at 95% with level of significance set at *P*< 0.05. Linkage disequilibrium (LD) was conducted with on-line software from HIV databases.

## RESULTS

### Sociodemographic Characteristics

The sociodemographic characteristics of the 271 couples by site of recruitment. Their ages ranged from 20 to 60 years with median age of 38 years. Over two-thirds were people of reproductive age group (31-40 years). Sixteen of the couples (3.0%), were engaged to be married, co-habited but were sero-discordant. The ethnicity showed that Ibos were more in the population of sero-discordant couples (44.1%) followed by the Yoruba (16.1%), only 9.3 % and 14.9% were of the Northern and other ethnic extractions. Overall, 224 (82.7%) were HIV sero-discordant; 26(9.6%) were concordant HIV positive and 21(7.7%) concordant HIV negative couples, as shown in Table 1.

### HLA-C Alleles Frequencies

Table 2 shows the frequencies of HLA-C alleles of high and low-resolutions in HIV positive and negative subjects. The highest frequency alleles observed in HIV negative subjects on low resolution were: C*04 124 (49.3%) followed by C*07 86(31.6%). In high resolution, C*04:01 106(39.0%) had the highest frequency, followed by C*07:01 64(23.5%). The least frequently encountered alleles were C* 01:02, C*12:03, and C*14:01, with each having frequency of 0.4%. The highest frequency alleles observed in HIV positive subjects were: C*04 92(39.0%) followed by C*07 81(34.3%) for low resolution. The highest frequency observed in the high-resolution alleles were C*04:01 72(34.5%), followed by C*07:01 57 (24.2%). The least occurring HLA-C alleles were: C*02:26 1(0.4%), C*03:03 1(0.4%), C*03:03 1(0.4%) and C*04:15 1 (0.4%).

**Table 2:**
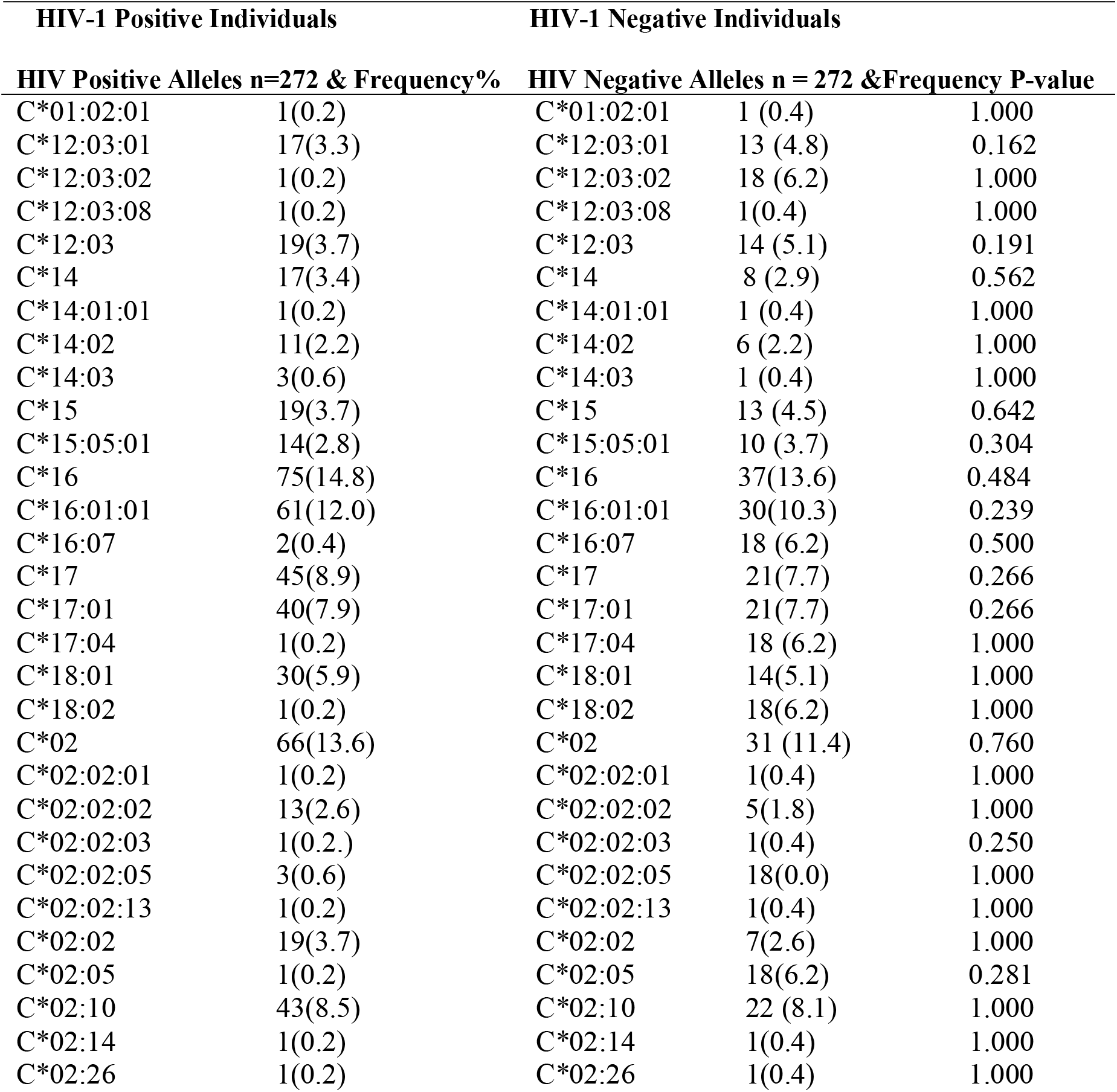
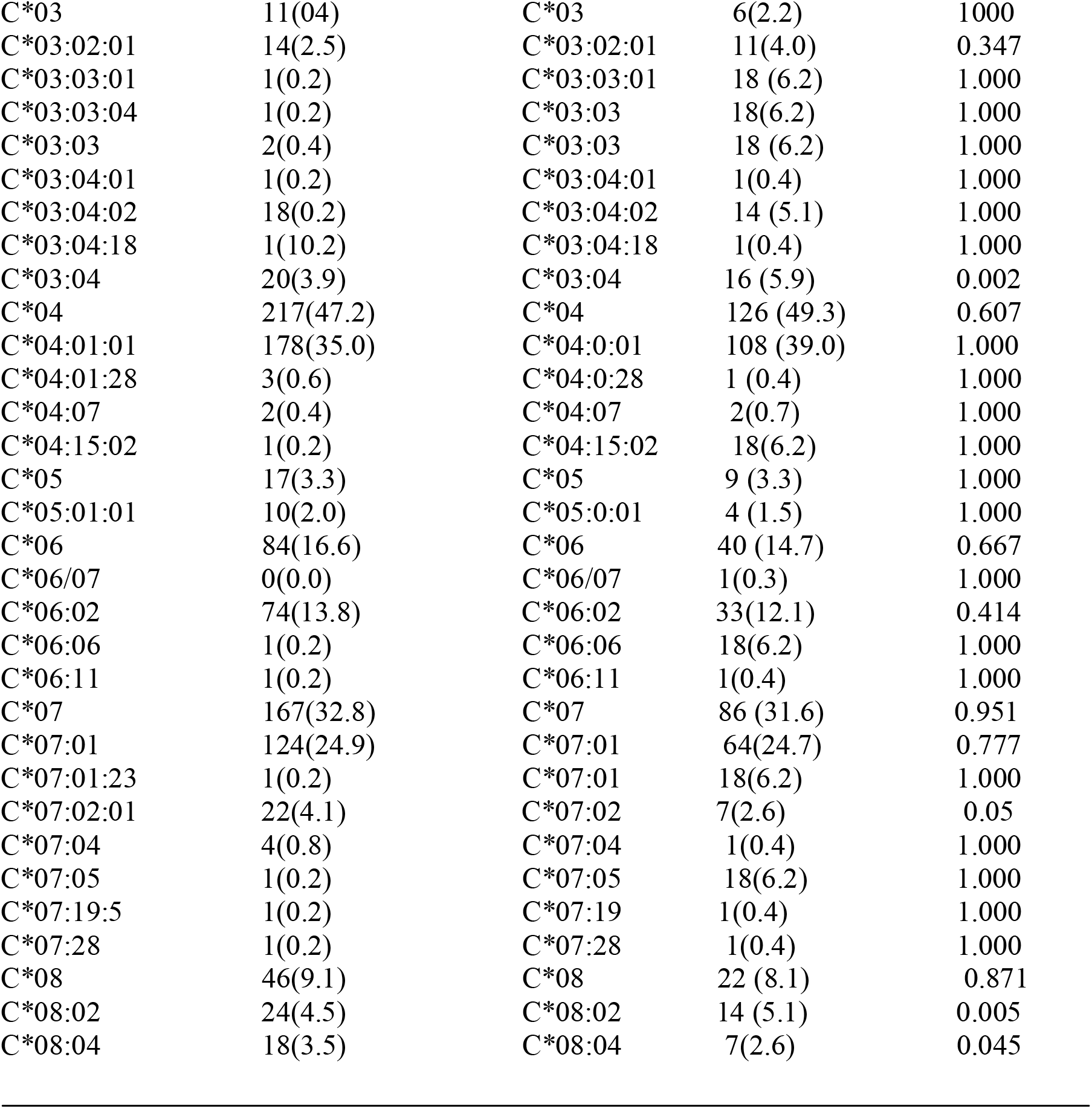
Frequencies and Cross tabulation of Major HLA-C Alleles and Haplotypes in HIV-1 Positive and Negative Individuals.

### Association of HLA-C and HIV-1 Susceptibility in Serodiscordant couples

Figure 1 shows the Chi-square analysis of HLA-C Alleles C*07:02 (OR = 4.19, *P<0*.*05*) and C*08:04 (OR = 3, *P*<0.045) which were found to be independently associated with HIV-1 susceptibility in the cohort.

**Figure 1:**
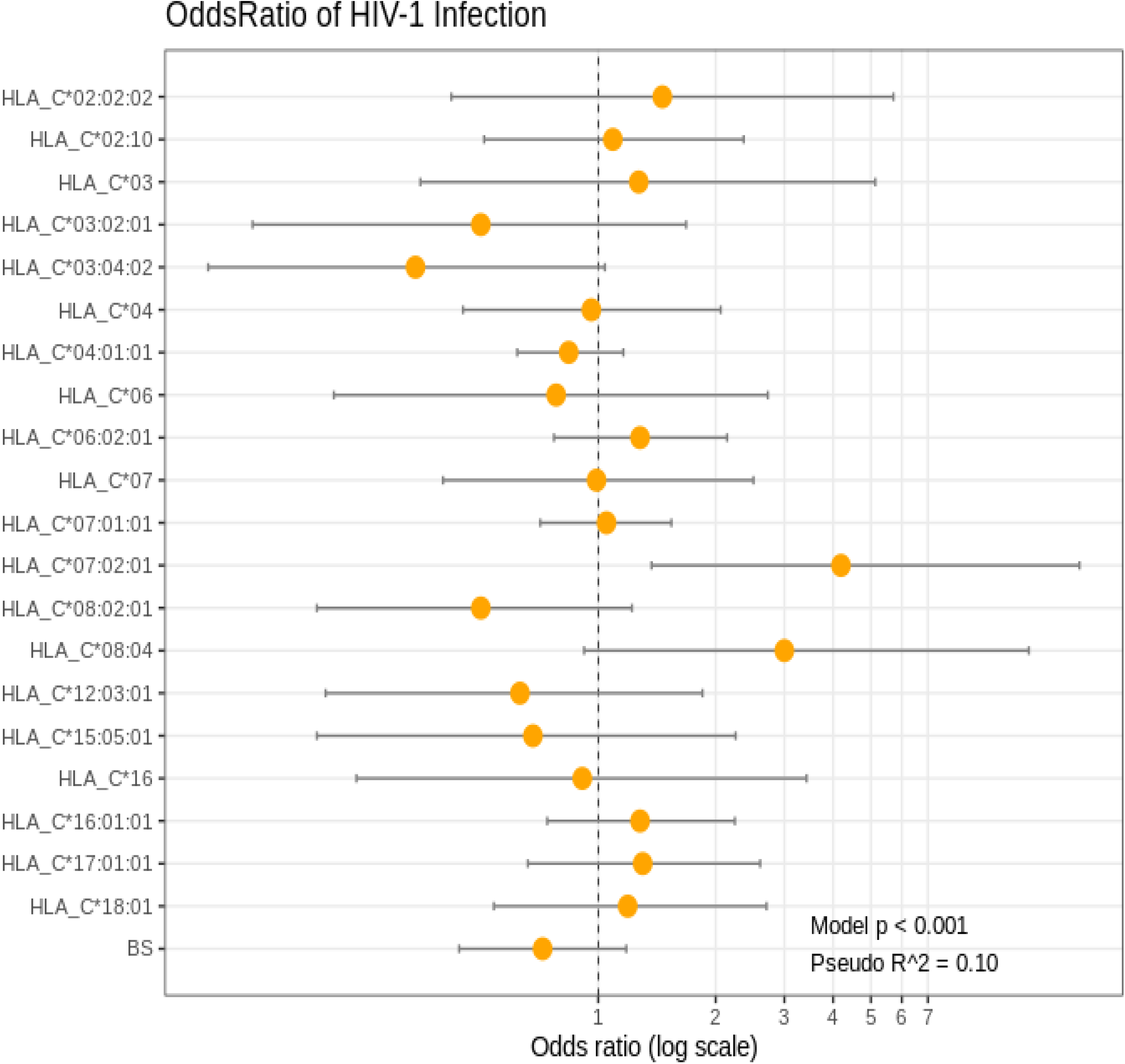
HLA-C genotypes Associated with Resistance and Susceptibility to HIV-1 Infection.

### Association of HLA-C and HIV-1 Resistance in Serodiscordant couples

HLA-C*08:02 (OR = 0.5, *P*<0.005) and C*03:04 (OR = 0.34, *P*<0.002) were significantly associated with HIV-1 resistance to HIV-1 infection.

### Long Term Non-progressors and their Partners

The longest period any of them have lived without drugs is > 7 years (12 HIV positive individuals) and the shortest is 0-12 months (57 HIV positive individuals). The profile of Long-term Non -Progressor’s (LTNP) among HIV positive individuals were reviewed along differential HLA types, their partners, baseline, follow-up of CD4+ count and plasma viral loads.

It was observed that this group of indexes have HLA-C genes associated with low plasma viral loads (HIV-1 RNA) and high CD4 T cell counts. However, their partners (80%) were HIV Negative. HIV susceptible alleles (C*07:02) was identified in 5 individuals and a couple inherited the allele C*07:20:01 in allele group level.

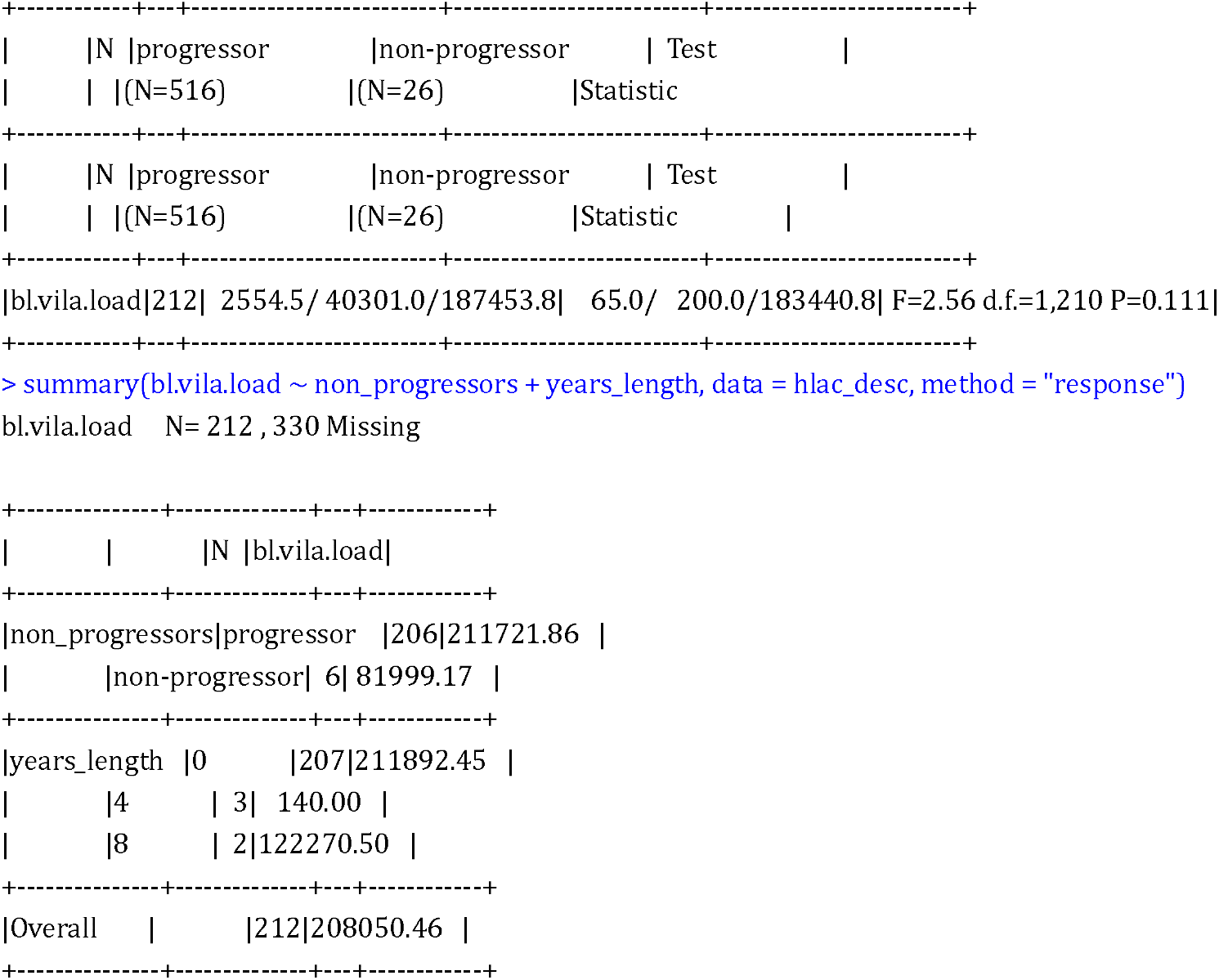

Analysis was done using R-programming language (R Core Team, 2020). Data was cleaned and prepared for analysis using Hmisc package for R-programming (Harrell, 2020). Bridging ImmunoGenomic Data-Analysis Workflow Gaps (BIGDAWG) was used to analyze polymorphic HLA-C data generated in the study to provide information on Hardy-Weinberg equilibrium, allele frequencies, odds ratios, confidence intervals and p-values for each allele (Pappas *et al*., 2016), R Core Team (2020) and Frank E Harrell.

## DISCUSSION

Previous studies reported that couples who do not have knowledge of their HIV status gave room to increased heterosexual HIV transmission among them in sub-Saharan Africa (Idoko *et al*., 2015, Allen *et al*., 2007 and Begala, 2006). Again, it has been reported that HIV-1 serodiscordant couples produced clear evidence that HIV-1 transmission from HIV positive (index) to seronegative partners depended on the specific HLA class 1 alleles which is also dependent on the strong plasma viral load (PVL) (HIV-1 RNA) and immunological control CD4 T cell count of the index (Peterson *et al*., 2015 and Tang *et al*., 2008). Therefore, a comprehensive analysis of HLA-C polymorphisms associated with HIV-1 transmission amongst heterosexual serodiscordant couples was investigated.

Our result in figure 1 showed that some HLA-C alleles have some protective and susceptible effects across the HLA-C class I locus which is also dependent on individuals’ plasma viral load and immunological control, CD4 T cell count. HLA-C*07:02:01 (OR = 4.19, *P<0*.*05*) and C*08:04 (OR = 3, *P*<0.045) were found to be independently associated with HIV-1 susceptibility or HIV diseases progression in the cohort of serodiscordant couples in Nigeria. HLA-C *07:01 alleles were significantly associated with high plasma viral loads (*P<0*.*004)* when compared with *08:04 that was moderately associated with high plasma viral loads (*P<0*.*06)*. HIV negative partners of the index case with these alleles (C*07:02:01 (OR = 4.19, *P<0*.*05*) and C*08:04 (OR = 3, *P*<0.045) are likely to seroconvert. With these alleles (C*07:02:01/C*08:04), HIV transmission to the HIV negative partner may be possible, if the index case is a male and have other clinical markers of HIV disease progression such as presence of untreated Sexually Transmitted Infections (STIs), other behavioural factors such as: age, non to inconsistent condom use, type of sexual activity involved in, multiple sexual partners and inconsistence in the use of ARVs. This result is consistent with a study done in Kenyan Pumwani Sex Workers’ cohort where alleles C*07:02:01/ B*07:02:01 were recorded as being associated with rapid HIV seroconversion as reported by Peterson *et al*. (2015). However, HLA-C*08:04 18(3.5%) is being reported for the first time to be associated with HIV seroconversion or HIV susceptibility in Nigeria. HLA-C*08:04 was identified in the HLA-C locus of African American population as reported by Tu *et al*., 2007 and so was consistent with West African population HLA specific diversity. This observation calls for further studies on the mechanism by which C*08:04 allele may influence HIV seroconversion in the cohort of serodiscordant couples. Previous studies by Bachtel et al., (2018) showed that HLA alleles that are not protective have affinity for Nef protein. Nef protein reportedly destroys HLA, CD4 +, and CD28+ cell surface expression and protect HIV infected cell from CTL recognition and destruction. By the destruction of HLA cell surface expression, there is down regulation of subset of HLA -C, -A, -B and E molecule which prevents HIV recognition by NK cells for destruction Blais *et al*., (2011).

We also observed that some individuals that have HLA-C*08:02 (OR=0.05, *P*<0.005) and C*03:04 (OR= 0.34, *P*<0.002) alleles were significantly associated with HIV-1 resistance. Majority of the individuals having HLA-C*03:04 were significantly associated with high CD4 T cell *(P= 0*.*002*) and baseline plasma viral loads ranging from 20-645copies/ml and so, are less likely to seroconvert. The result from this study is comparable to some reports from previous studies where the partner of the index may remain seronegative if the HIV-1 RNA of the index is consistently at 20copies/mm3 (lower limit of detection of either 40 or 50 copies/ml) as reported by Aidsmap, (2016). It is also comparable with reports from Quinn *et al*., (2000) who established from their studies that, if the HIV-RNA remains at 20copies/mm3, the index may not be able to transmit HIV to his/her negative partner. Furthermore, having an undetectable plasma viral load (HIV-1 RNA) confirms that the risk of HIV becoming resistant to ARV is very minimal. This further shows that, with improved immune system, the risk of having opportunistic infections is reduced. Though HLA-C*08:02 and C*03:04 were newly observed to show resistance to HIV −1 acquisition in Nigeria, it is possible that their cytotoxic lymphocyte expression improved HIV control through active cytotoxic CD8^+^ T cell responses. (CD8**+** cytotoxic T lymphocytes CD8^+^ T directly killing virally infected cells and producing antiviral cytokines).

Some HLA-C alleles identified in Kenyan studies (Peterson *et al*., 2015) as protective alleles include: C*06:02 and C*07:01 which is contrary to our findings. Our study observed that they were associated with very high plasma viral loads and low CD4^+^ T cells *(p =*0.006 and *p* = 0.003). Borghans *et al*. (2007) in their study showed that HLA alleles that is, defending the immune system have a true preference for the p24 Gag protein and CTL responses against Gag which moderately reduces HIV disease evolution.

The Long-term Non-Progressor’s (LTNP) HLA-C locus contains differential allele types which fall among *04:01:01 (50%), 17:01 (40%) and 07:01(30%) in Nigeria. The reason for this preferential HLA types is not well understood so can be further investigated. Another study that is consistent with our study, identified C*07 and C*04 restricted Cytotoxic Lymphocytes in Long Time Non-progressors. However, Zepeto and Beretta, (2012) identified C*07, C*04 and C*17 as inhibited alleles or at-risk alleles that are not protective. This is because they contain an intact miR-168 biding site and their expression on the surface are down -regulated. Our study further observed that this group of indexes (LTNPs) has HLA-C genes associated with low plasma viral loads (HIV-1 RNA) and high CD4 T cell counts. Davis (2014) recent study, reported in his study that 11 of the 13 LTNP (85%) had a gene encoding an HLA variant called HLA-B*5701 which has been implicated to slow HIV disease evolution. However, in this study, HLA-C gene was found restricting HIV-1 multiplication in 6.2% of 271 serodiscordant couples and delay AIDS on-set in these indexes. In another previous study, LTNPs was associated with a weak or non-active form of HIV, but it is now known that many LTNP patients carry a virulent form of the virus HLA-C O’connell *et al*., (2009) and Blankson *et al*., (2009). HLA-C LTNPs may be associated with increased surface expression which is associated with reduced viral load and low CD4^+^ T cell counts, or the alleles have preference for p24 Gag protein which reduces HIV diseases progression. This High HLA-C expression have improved HIV control through active cytotoxic CD8^+^ T cell responses as recorded in African and European Americans (Apps *et al*., 2013).

High resolution frequencies observed in HIV-1 positive individuals were: C*04:01:01 (178, 24.7%) and C*07:01 (124, 24.9%) compared to HIV negative individuals: C*04:01:01 (108, 39.0%) followed by C*07:01 (64, 24.7%). These results showed consistency with a study done in Kenyan where C*07:01 (16.1%) and C*04:01:01 (12.5%) had the highest frequencies in their study population, (Peterson *et al*. 2015). Interestingly, one of the rarest alleles, C*01:02:01, identified in this study: (1, 0.2% HIV +) and (1, 0.4% HIV-), was also rarely observed among Kenyan population. In Harare Zimbabwe also, C*04:01:01 (25.2%) was the most frequent allele observed in the population of 123 children and adolescent aged 10-18 years Ferrand *et al*., (2015) which is also consistent with this study. Peterson *et al*., (2014) reported the diversity and frequencies of HLA-alleles and haplotypes among the East African Population C*06:02:01(31.37%), C*04:01:01(31%) and C*07:01:01(26.35%) which is also consistent with our findings. This is showing that there are similarities in HLA -C haplotypes and alleles of all African descent.

## CONCLUSION

This study has shown how some HLA alleles (C*07:02:01 /C*0804) and (C*08:08/C*03:04) contributed to HIV-1 disease susceptibility and resistance in serodiscordant couples in Nigeria. The mechanism involved CD4^+^ T helper cells and cytotoxic lymphocytes immune responses that control the spread of chronic and acute HIV-1 diseases. This information may contribute to the development of future effective HIV vaccine in Nigeria. It was also discovered that C*04:01:01 and C*07:01 has the highest frequency distribution in both HIV positive and HIV negative individuals. This is showing that there are similarities in HLA -C haplotypes and alleles of all African descent.

## SUMMARY SENTENCE

a. Some people living with HIV-1 (index) can transmit HIV-1 to their negative partners through sexual exposure. This is because these partners have HLA-C*070201 (OR = 4.19, *P<0*.*05*) and C*0804 (OR = 3, *P*<0.045) which is susceptible to HIV-1 transmission.
b. Furthermore, some partners who have favourable HLA-HLA-C*0802 (OR=0.5. *P*<0.005) and C*0304 (OR=0.34. *P*<0.002) alleles have been identified to protect them from contracting HIV-1 from their index.
c. The highest frequency distributions of high-resolution HLA-C alleles observed in people living with HIV-1 were: HLA-C*040101 178(35.0%) and their partners C*040101 108(39.0%).

## Data Availability

Besides those presented in the study, there are no other supporting data for this manuscript

## ETHICAL APPROVAL AND CONSENT TO PARTICIPATE

Ethical approval was sought and obtained from NIMR IRB (IRB/12/176) and NAUTH Ethics Committee (CS/66/7/79). Confidentiality was duly maintained, although necessary information about the participants were disclosed only to the clinicians managing the participants – in the various clinics. Only consenting participants were enrolled.

## CONSENT FOR PUBLICATION

The participants were aware and provided informed consents to participate and to publish results of the study in academic journals.

## AVAILABILITY OF SUPPORTING DATA

Besides those presented in the study, there are no other supporting data for this manuscript.

## COMPETING INTERESTS

The authors report no competing (commercial/academic) interests.

## FUNDING

Funding for this study was provided by the first author, Dr. Ma Lou, HIV Research Trust Scholarship UK and Prof. Babatunde Salako, the Director General of the Nigerian Institute of Medical Research.

## AUTHORS’ CONTRIBUTIONS

NMO was involved in the conceptualization and design of the study, collected sample from first site and wrote the paper.

ML worked on the HLA analysis and sponsored the genomic study.

OIE and NNO supervised the project and proofread the manuscript.

FE and PE collected and processed the samples at the second site.

AJ analysed the data and edited the manuscript.

TB helped with running the PCR. MMO edited the manuscript.

All authors read and approved the final version of the manuscript.

## ACKNOWLEDGEMENTS

**I wish to acknowledge:**

1. HIV Research Trust Scholarship UK 2012 (HIVRT12-082)
2. Dr. Ma Lou. Associate Professor, University of Manitoba, HIV Host Genetics, National Microbiology Laboratories Winnipeg Canada (sponsorship on genomic studies).
3. Prof. Babatunde Salako, the Director General of the Nigerian Institute of Medical Research: Pre-Doctoral Research Development Grant

## Notes

### Competing Interest Statement

The authors have declared no competing interest.

### Author Declarations

Ethical approval was sought and obtained from NIMR IRB (IRB/12/176) and NAUTH Ethics Committee (CS/66/7/79).

